# A Heuristic Model for Spreading of COVID 19 in Singapore

**DOI:** 10.1101/2020.04.15.20067264

**Authors:** Fook Hou Lee

## Abstract

This paper presents a simple heuristic model for COVID 19 spreading. The model is based on a propagation unit of time. The state of the epidemic at the end of the time unit is then related to that at the start through recurrence relationships. By propagating these relationships over the required number of time units, a projection can be made over time. The model is readily implemented on a spreadsheet and is therefore potentially widely accessible. It can serve as a useful tool for scenario planning and forecasting not just for an entire population, but also for a specific community within a population.

## 1 Introduction

The COVID 19 pandemic has challenged a lot of our conventional notions derived from previous experience. Many questions regarding this pandemic remain unanswered to date and some of these relates to the manner in which the epidemic unfolds and the strategies of controlling the epidemic. Much of the work to date relates to characterising the virus, development of testing technology, cures and vaccines. Some models have also been advanced to simulate the spread of the disease. Fang et al. (2020) modelled the transmission dynamics of COVID 19 using a variation of Kermack and McKendrick’s (1927) SIR model. Yeghikyan (2020) also used Wesolowski et al.’s (2014) variation of the SIR model which took into account mobility patterns. Other forecasts have also been made by research organisations, using mainly data-driven softwares (e.g. IHME 2020; Adam 2020). Many of these are based on the SIR approach, but implemented in highly proprietary frameworks which are not readily accessible to third parties. Other approaches have also been used. For instance, Sanche et al. (2020) proposed that, if the basic reproduction number R_0_ is 6.7, then even “*if as low as 20% of infected persons are asymptomatic and can transmit the virus, then even 95% quarantine efficacy will not be able to contain the virus*”. However, Sanche et al.’s (2020) findings were based on the basic theory on the force of infection, rather than a full simulation and focused on the asymptomatic cases. Hence, it is a partial model. Furthermore, current understanding appear to suggest that infectivity is probably much higher amongst the symptomatic cases, especially within the first three days of symptoms onset, compared to the asymptomatic case (WHO 2020). Kermack and McKendrick’s (1927) original SIR model was expressed in closed-form mathematical relationships. Individual researchers are now much more empowered in terms of computing facilities than their predecessors in Kermack and McKendrick’s era. There is thus a case for the development of models which are readily implemented on personal computers, but at the same time, sufficiently reliable in their prediction.

This paper presents a heuristic epidemic spreading model, which can be readily implemented, modified, up-scaled and generalised personal computer spreadsheet. The model based on two parameters, namely detection efficiency and daily infectivity. The model formulation is first discussed. Examples are then presented to verify and calibrate the parameters, with respect to COVID 19. Some findings from the simulation, relevant to COVID 19, particularly for the Singapore context, are highlighted. Finally, an extension of the model is also made to include the effects of herd immunity and asymptomatic period.

## 2 A Simple Framework

The proposed model is akin to the SIR model, in that both are process-based. However, this model is formulated in discrete format, wherein the starting point is a propagation unit, taken herein to be of a duration of 1 day. Fig. 1 illustrates the propagation process of this model. At the start of day 0, an assumed number of active infected cases is seeded into the population. During the course of the day, detection processes will identify and remove a certain fraction, designated by *k*, from the infected cases. The remaining undetected cases will then go on to re-infected other cases, based on a daily infectivity ratio *r*. The typical sequence of operations for three days is shown in Fig. 1, wherein the variables are defined as follows:

**Fig. 1.**
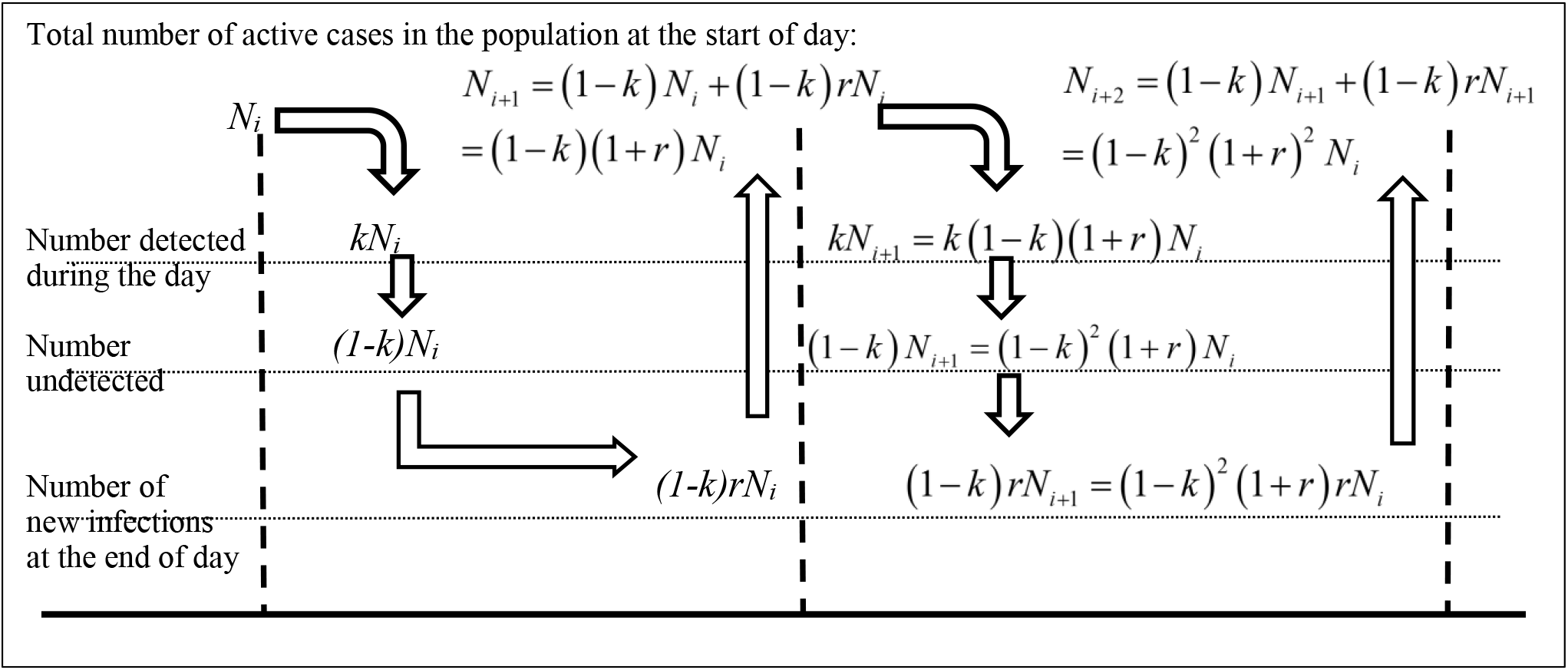
Propagation process of proposed model.

*N*_*i*_, *N*_*i*+*1*_ and *N*_*i*+*2*_ = the active number of infected cases still in the population at the start of the three respective days i.e. *i, i*+*1 and i*+*2*. This does not include those cases which have been previously detected and removed from the infected reservoir. Hence, it is not the same as the total number of infected cases.

*k* = the detection efficiency. *k* = 1 means 100% detection, i.e. every infected case is detected. *r* = the average daily infectivity of each undetected infected case, e.g. *r* = 2 means every undetected case went on to infect another 2 persons within a day.

As the arrows highlighting the sequence of operations in Fig. 1 show, the number detected during the course of each day is subtracted from the number at the start of the day. The undetected number is then allowed to re-infect new cases which, together with the undetected cases, goes on to form the active cases at the start of the following day. For simple scenarios, mathematical relationships for this recurrence model can be derived, as shown in the sections below.

## 3 Total Number of Reported Case (M_j_)

The total number of case that is usually reported M <u>**is actually the cumulative number of cases detected**</u>. Summing over j days,

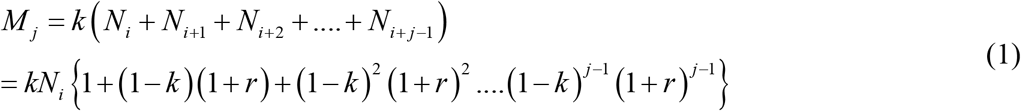

If *k* and *r* are constants, then Eq. 1 is a geometric series, which, when summed to the *j*^*th*^ term gives

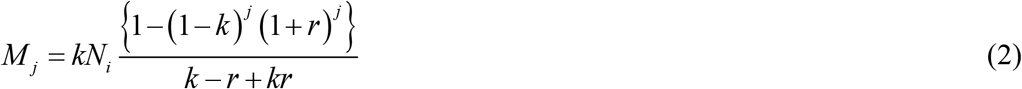

We note that M_j_ increases monotonically with *j*, i.e. the number of days.

## 4 Number of undetected infected persons

*M*_*j*_ does not relate directly to the probability of encountering an infected person at any point in the population. This probability is represented more by the active number of undetected infected persons on a given day, say the *j*^*th*^ day, *P*_*j*_, which is given by

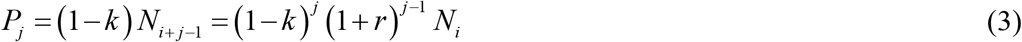

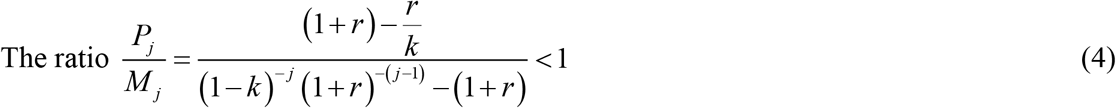

and decrease as j increases. Hence, the total number of reported cases is not representative of the actual probability of encountering an infected person in the population.

## 5 Number of active, detected and undetected cases

The number of active cases at the start of the *j*^*th*^ day is *N*_*j*_. For this number not to increase, *N*_*i*+*2*_ *≤ N*_*i*+*1*_ *≤ N*_*i*_*…*..*≤ …* .*N*_*j*_, which would require that

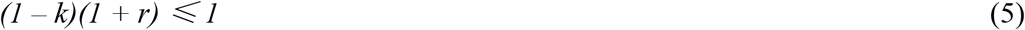

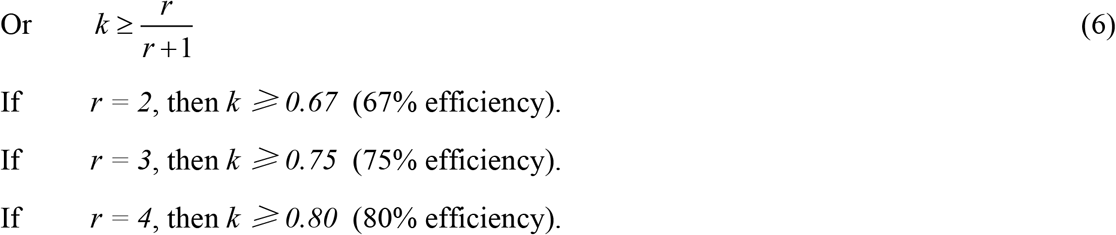

Hence, to decrease the active number of infection day-by-day, the detection efficiency has to be sufficiently high in relation the daily infectivity. As discussed below, the latter is a function of the disease, infectious period, community and social setting. For instance, a disease in a setting where people are isolated may be a low *r*-value. The same disease in a setting where people are highly socialised would have a higher *r*-value.

It should be noted that the *N*-value is not the number of new cases detected each day. That number *Q*_*j*_ is given by *Q*_*j*_ *= kN*_*j*_. However, the criterion for stabilization is similar, if the number detected each day is to remain steady or decrease, then

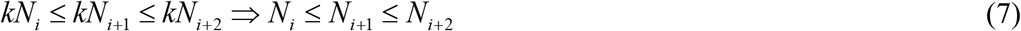

which also requires Eq. 6 to be satisfied. It should be noted that if the incremental number of reported cases is steady day-upon-day, then

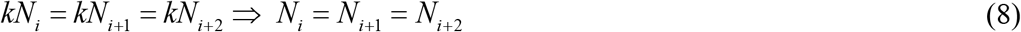

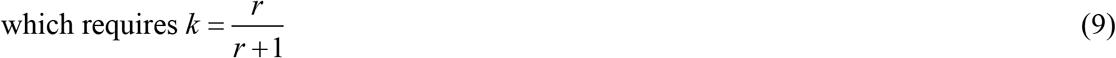

Hence, Eq. 9 is the condition for the curve of cumulative reported number of case to reach the point of inflexion. At this point inflexion, the rate of detection is just sufficient to offset the rate of new infection. At this point, the battle is not won, but it is also not lost. It is akin to a stalemate situation.

The undetected cases in each day *P*_*j*_ is given by (1− *k*) *N j* etc.. and can be related to *Q*_*j*_ by

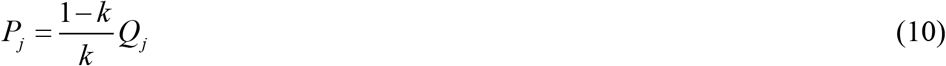

The number of undetected cases *P*_*j*_ affects the probability of encountering an infected person in the population. If *k* can be estimated, then *P*_*j*_ can be readily estimated from *Q*_*j*_.

To illustrate the significance of this, if the number of detected cases on a certain day *Q*_*j*_ = 120 (this being the peak number in Singapore, on 5 April 2020) and the detection efficiency *k* = 0.5, then number of undetected cases *P*_*j*_ *= 120*. If the daily infectivity *r* = 2, then at the start of the following day, we would expect the number of active cases to be ∼360 and an upper-bound estimate of the probability of encountering an infected person in Singapore 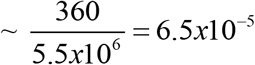 or roughly 1 in 15000, based on a population size of 5.5 million. In other words, one can, on average, expected to encounter an infected person in every 15000 encounters. On the other hand, using the cumulative number of detected cases would give a probability of 235 per million population (https://www.worldometers.info/coronavirus/#countries), or 1 in 4200, which is ∼3.6 times higher. Hence, the cumulative number of detected cases is not directly related to the chances of encountering an infected person. On the other hand, if *k* = 0.1, then *P*_*j*_ *= 9Q*_*j*_ and every detected cases implies 9 undetected cases. In other words, the reservoir of undetected active cases will far exceed the number of detected cases and the latter ceases to be meaningful.

## 6 Programming the framework

The mathematical results presented above only applies to very simple scenarios. However, the recurrence relationships can be readily upscaled and generalised to more complex scenarios using a spreadsheet or other programming environments. The examples below were calculated by incorporating these recurrence relationships into a Excel spreadsheet.

### 6.1 Examples of application

#### 6.1.1 Constant k and r

We now consider the simplest case where the daily detection efficiency k and infectivity r are both constant. Figs. 2a and b shows the effect of different values of *k* for *r = 2*. As can be seen, the trend is highly sensitive to the detection efficiency and daily infectivity. A slight drop in k or a slight increase in *r* is enough to cause a drastic increase in the numbers of cumulative and incremental detected cases. The case of k=0.6 in Fig. 2a also shows what happens in a yet-to-stabilised epidemic situation. As can be seen, the upper part of this curve is almost linear, indicating a log-linear relationship between *M*_*j*_ and number of days *N*, of the form

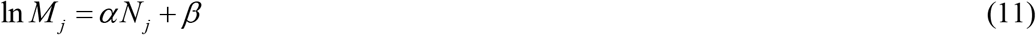

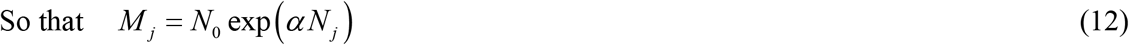

**Fig. 2.**
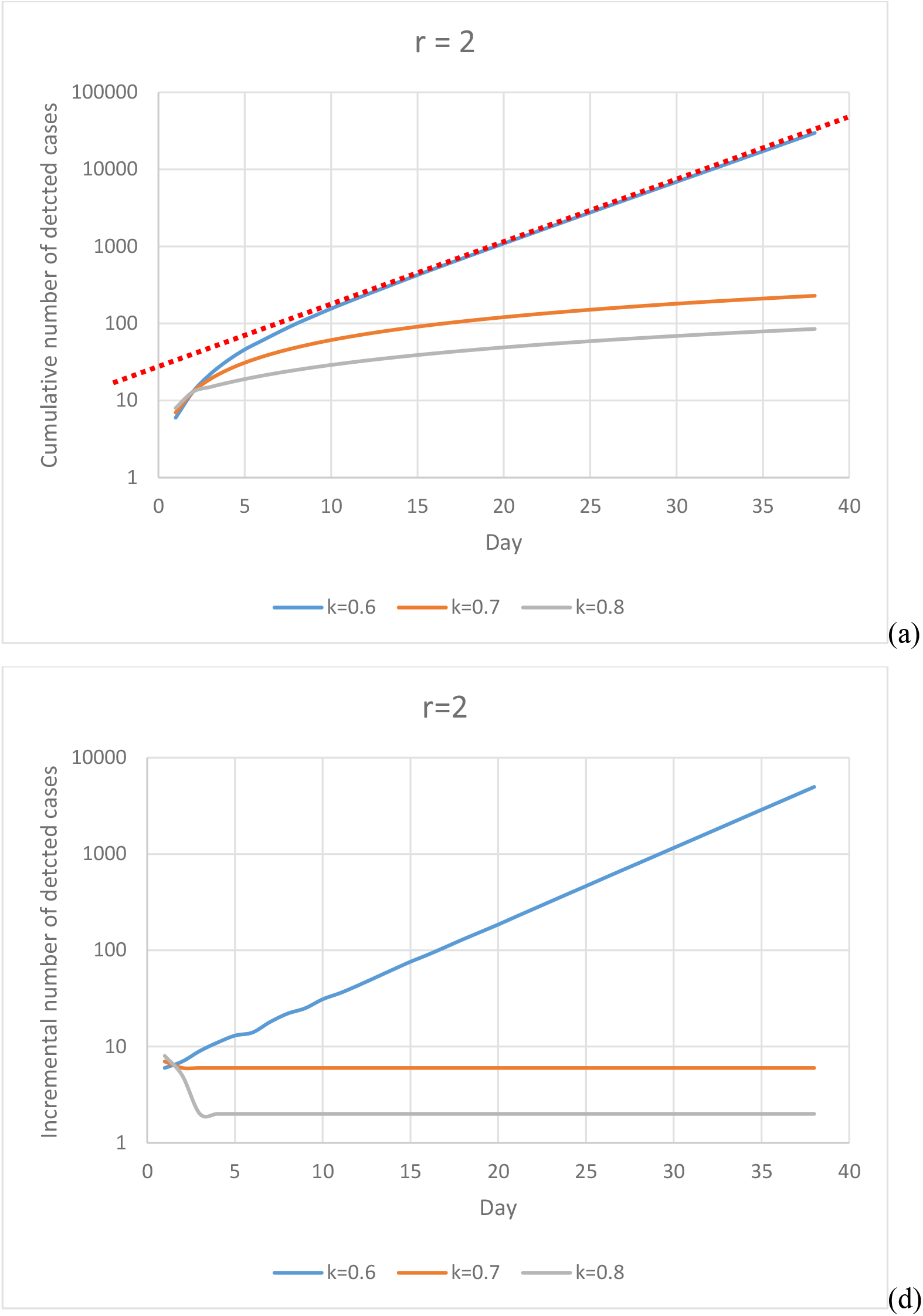
(a) and (b). Cumulative and incremental number of detected cases for k ranging from 0.6 to 0.8 and r = 2.

In which *N*_*0*_ *= exp(β)* is the initial number of infected persons (at 0^th^ day). The parameter *α* governs the rate of exponential increase and is dependent upon *k* and *r*. The effect of α can be assessed by considering the value of *M*_*j*_ at intervals of a specified number of days *ΔN*. For instance, on the *j*^*th*^ day,

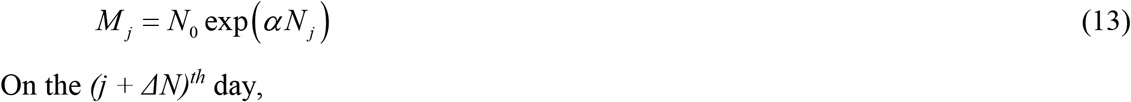

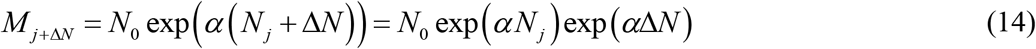

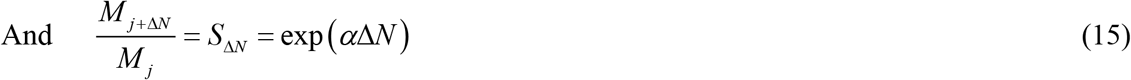

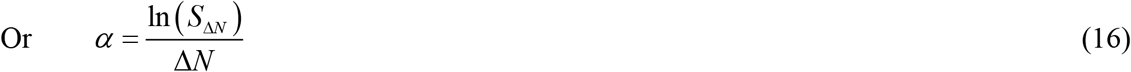

If *S*_*ΔN*_ is chosen to be 2 and *ΔN*_*2*_ is the number of days for a doubling of the detected cases, then

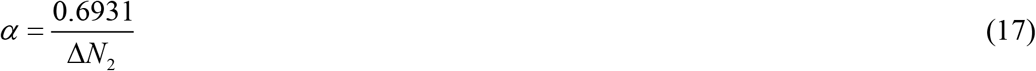

This is consistent with the exponential growth of an epidemic. This log-linear trend is also demonstrated in the early-day trends in several countries, Fig. 3. This indicates the projection results are consistent with conventional wisdom as well as data of COVID 19 cases. For instance, in Fig. 3, initial Chinese data show a doubling once every ∼1.5 days. Substituting S_ΔN_ = 2 and ΔN = 1.5 gives α = 0.462. The US data show a doubling once every ∼2.5days, indicating that *α* = 0.277. This motivates the possibility of back-calibrating the value of *k* through *α*, if *r* can be independently estimated. This is further discussed below.

**Fig. 3.**
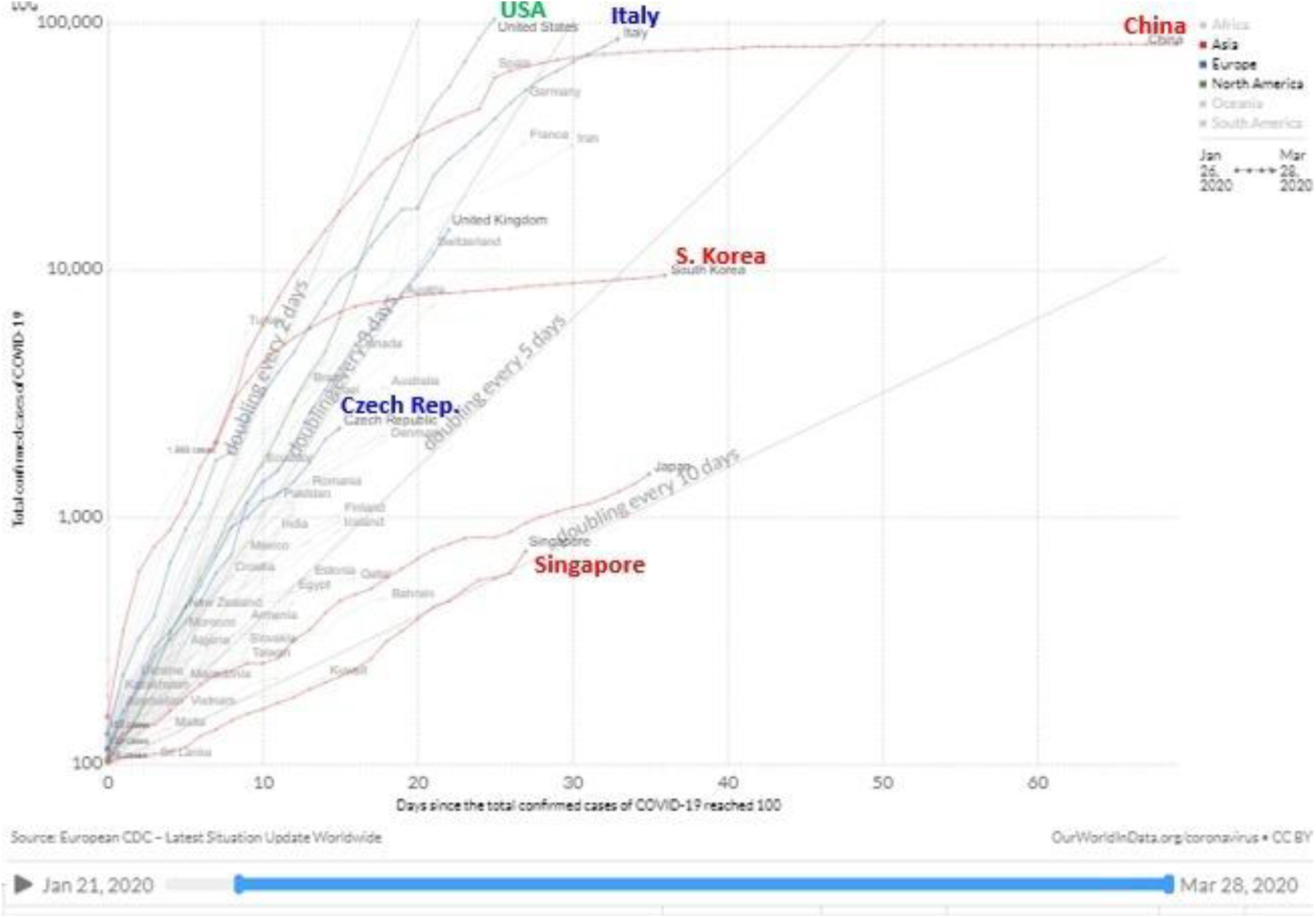
Total number of reported coronavirus cases in various countries (after https://www.visualcapitalist.com/infection-trajectory-flattening-the-covid19-curve/).

As Figs. 4a – c show, the ratio 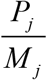 decreases monotonically in all cases. In scenarios which have yet to stabilise e.g. Fig. 4c for 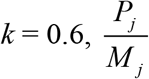 stagnates at a relatively high value, but which is, nonetheless, still significantly less than 1, indicating that the cumulative detected cases may not serve as a good representative of the number of active cases.

**Fig. 4.**
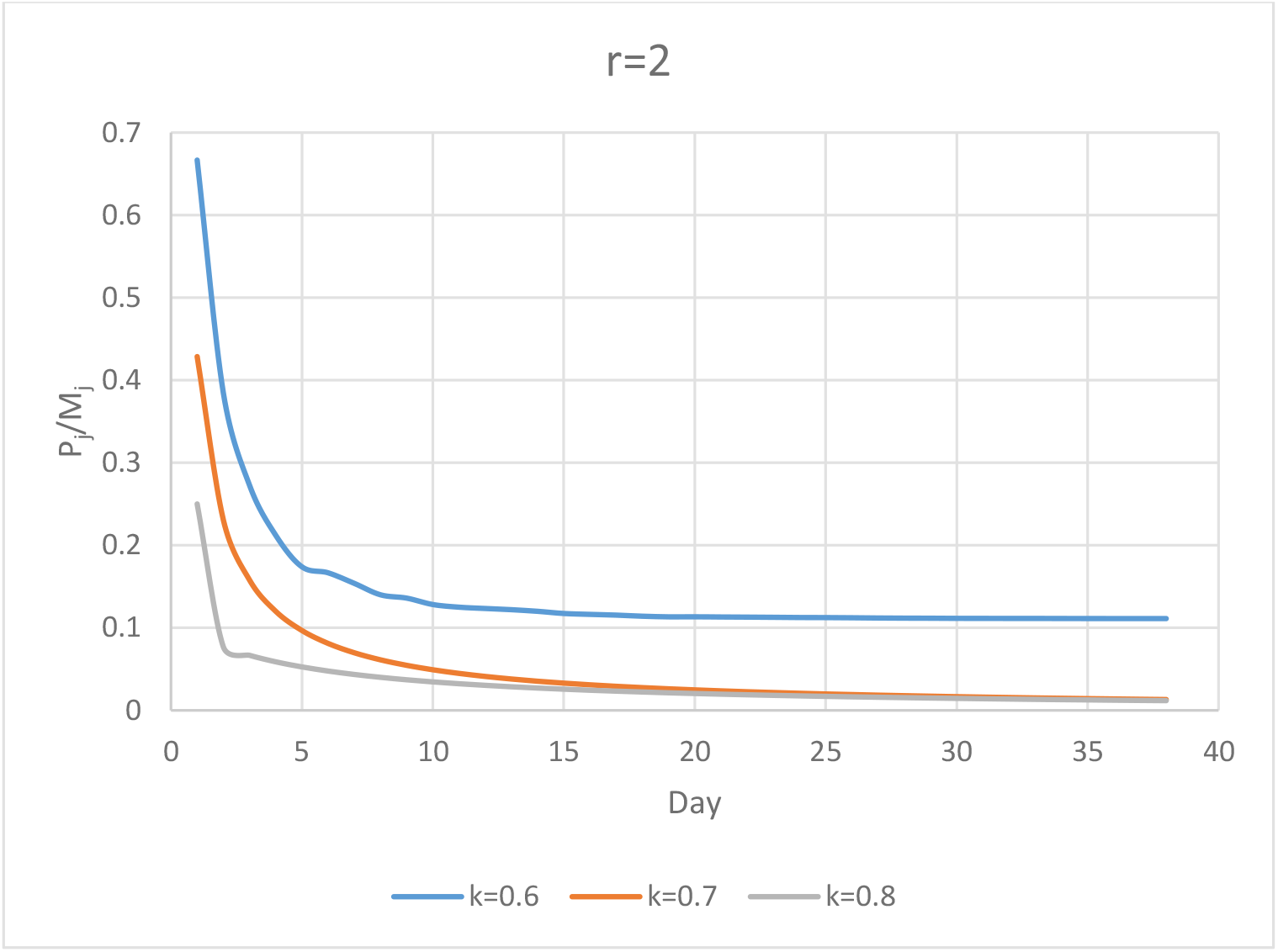
Evolution of 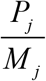 for different *k* and *r*-values.

#### 6.1.2 Variable k, constant r

We now consider a case where the detection efficiency starts at a lower level but was ramped up at different points and with different rates, using the following strategies:

a. *k* = 0.6 up to the 10^th^ day and then ramped up to 0.9 on the 16^th^ day.
b. *k* = 0.6 up to the 15^th^ day and then ramped up to 0.9 on the 30^th^ day.
c. *k* = 0.5 up to the 15^th^ day and then ramped up to 0.9 on the 35^th^ day.

As Fig. 5a shows, the initial exponential followed by inflexion and then stabilisation is now clearly reflected. Furthermore, as Fig. 5b shows, the peaking of the incremental number of detected cases is also reflected in all three cases but is more pronounced in Strategy 3 owing to the later and slower ramp-up. Finally, the effect of increasing the *k*-value is also reflected in a decrease in 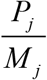.

**Fig. 5.**
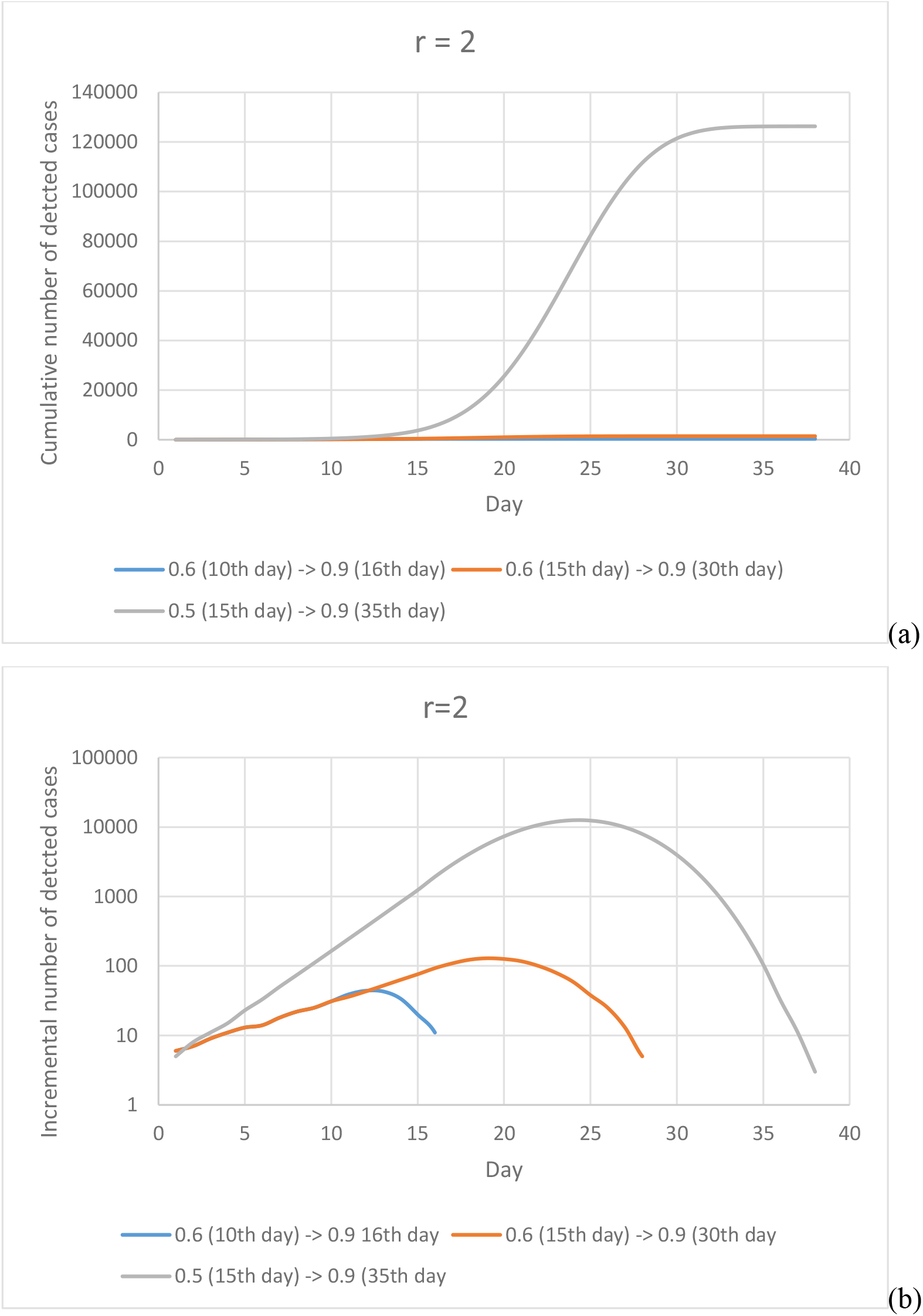
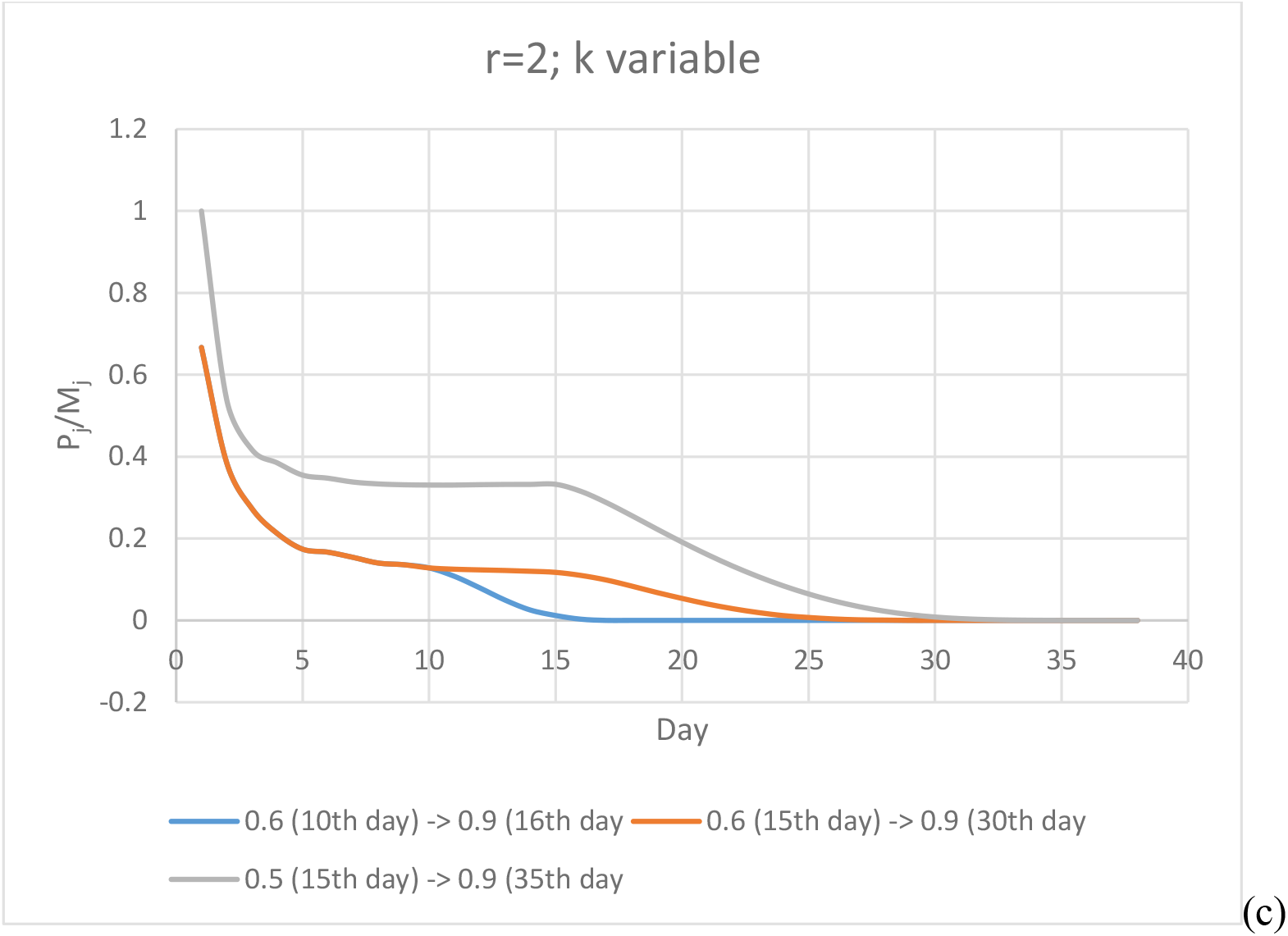
Results for *r* = 2 and variable *k*. (a) Cumulative number of detected cases (b) Incremental number of detected cases (c) 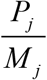.

## 7 The nature of the detection efficiency *k* and daily infectivity *r*

The daily infectivity *r* is similar, but not identical to the basic reproduction number *R*_*0*_ widely used in epidemiology (https://www.healthknowledge.org.uk/public-health-textbook/research-methods/1a-epidemiology/epidemic-theory). The basic reproduction number R_0_ is defined as “*the average number of secondary infections produced by a typical case of an infection in a population where everyone is susceptible*”. This ignores the effect of herd immunity. To account for herd immunity, an effective reproductive number R is also defined, such that

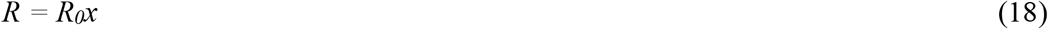

Where *x* is the fraction of the population that is still susceptible. Since R_0_ is the reproduction number over the infectious period of an infected individual and r is a daily rate

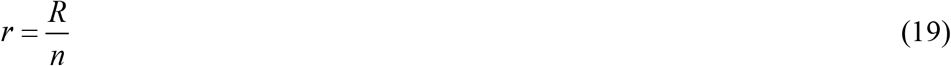

where *n* = the number of days over which infection can occur. If herd immunity is insignificant, then *R ∼ R*_*0*_.

It is well-established that *R*_*0*_ is not just dependent upon the infecting agent or the disease, it is also dependent upon a host of other factors including

- The rate of contacts in the community. This depends upon the social setting, proximity between people and whether they are using protective equipment and practising good personal hygiene etc. For instance, Zhang et al. (2020), using early stage data from the Diamond Princess cruise ship, estimated the *R*_*0*_ to range from 2.06–2.52, with a median at 2.28. Li et al. (2020) also estimated a similar range for *R*_*0*_, from 2.2 to 2.7. Leung et al. (2020) also estimated an *R*_*0*_ = 2.68. On the other hand, Sanche et al. (2020) estimated a much higher *R*_*0*_ ranging from 4.7 to 6.6. These differences may be attributed to different social settings and data accuracy. There may also be issues relating to undetected cases which may impact the estimated *R*_*0*_. Niehus et al. (2020), for instance, postulated that “…*the global ability to detect imported cases is 38% (95% HPDI 22% - 64%) of Singapore’s capacity*”.
- The infectious period. According to WHO (2020), “…*Data from clinical and virologic studies that have collected repeated biological samples from confirmed patients provide evidence that shedding of the COVID-19 virus is highest in upper respiratory tract (nose and throat) early in the course of the disease. That is, within the first 3 days from onset of symptoms. Preliminary data suggests that people may be more contagious around the time of symptom onset as compared to later on in the disease…….The incubation period for COVID-19, which is the time between exposure to the virus (becoming infected) and symptom onset, is on average 5-6 days, however can be up to 14 days. During this period, also known as the “presymptomatic” period, some infected persons can be contagious. Therefore, transmission from a pre-symptomatic case can occur before symptom onset. In a small number of case reports and studies, pre-symptomatic transmission has been documented through contact tracing efforts and enhanced investigation of clusters of confirmed cases. This is supported by data suggesting that some people can test positive for COVID-19 from 1-3 days before they develop symptoms….Thus, it is possible that people infected with COVID-19 could transmit the virus before significant symptoms develop. It is important to recognize that pre-symptomatic transmission still requires the virus to be spread via infectious droplets or through touching contaminated surfaces……An asymptomatic laboratory-confirmed case is a person infected with COVID-19 who does not develop symptoms….Asymptomatic transmission refers to transmission of the virus from a person, who does not develop symptoms. There are few reports of laboratory-confirmed cases who are truly asymptomatic, and to date, there has been no documented asymptomatic transmission. This does not exclude the possibility that it may occur. Asymptomatic cases have been reported as part of contact tracing efforts in some countries*”.
- The probability of infection being transmitted during contact, which is largely a characteristics of the disease.

Based on this, one can surmise that the infected person is likely to highly contagious within the first 3 days of onset of symptoms. Some transmission may occur during the asymptomatic period but does not appear to be as significant. It is also likely that after the first 3 days, he would have sought medical attention and would likely have been detected. Hence, it is postulated herein that an average estimate of the infectious period *n* ∼ 4 to 5 days. Together with the upper bound value of *R*_*0*_ of 6.6, this would imply *r* ∼ 1.3 – ∼1.7. However, this is an average figure; for special social setting, *r* may be much higher.

Less is known about the detection efficiency *k*. Niehus et al.’s (2020) postulation only relates to the relative (rather than absolute) detection capacity and they noted that Singapore’s detection capacity is likely to be less than 100% (or 1.0) and some other countries may be much lower. One way to back-estimate *k* is through the geometric rate of increase. If we assume a median value of *r* ∼1.5, based on the discussion above, then *k* can be estimated by back-fitting the *α*-value, Table 1. As Table 1 shows, Singapore’s back-fitted detection efficiency k is significantly higher than that of China (in the early stage) and the US. This is consistent with Niehus et al.’s (2020) postulated that “…*the global ability to detect imported cases is 38% (95% HPDI 22% - 64%) of Singapore’s capacity”*. The current back-fitted results indicate a somewhat higher percentage (53%) instead of 38%. Nonetheless, the trend is generally correct and it explains the slower rise in infected cases in Singapore. Moreover, the detection rate of 35.5% for China is also reasonably consistent with Wang et al.’s (2020) finding that “*at least 59% of infected cases were unascertained in Wuhan*”. The latter would imply a detection efficiency of 41% or less.

**Table 1.** Back-fitted detection efficiency k.

| Country | Number of days for doubling of detected cases | Measured $\alpha$ value | Best-fit $\alpha$ value | k-value for best-fit $\alpha$ value (r-value) |
| --- | --- | --- | --- | --- |
| US | 2.5 | 0.277 | 0.276 | 0.48 (1.5) |
| China (early stage data) | 1.5 | 0.462 | 0.461 | 0.355 (1.5) |
| Singapore | 10 | 0.0693 | 0.069 (curve is sub-exponential, showing that the epidemic was stabilising. Value was fitted to a chord on the curve). | 0.67 (1.5) |
| Singapore (14/3 – 9/4 Unlinked cases) |  | 0.141 | 0.14 | 0.54 (1.5) |
| Singapore (14/3 – 9/4 Unlinked cases) |  | 0.141 | 0.14 | 0.54 (1.5) |
| Singapore (14/3 – 9/4 Dormitory linked cases) |  | 0.376 | 0. | 0.54 (2.2) |

Fig. 6 shows the cumulative number of cases from dormitory clusters, non-dormitory clusters and unlinked cases from 30 March 2020 till 9 April 2020. As can be seen, both the non-dormitory clusters and unlinked cases plot to almost the same gradient, implying almost equal *α*-value, which can be fitted using *k* = 0.54. This suggests a possible decrease in the detection efficiency from the initial high value of 0.67. The alternative explanation is an increase in *r*-value, but this seems rather unlikely since the social setting, virological characteristics and infectious period have not changed. However, there may be some over-estimation of the unlinked cases as some of these might have been subsequently linked to existing clusters or cases. The dormitory clusters show a much higher *α*-value. Since the *k*-value is unlikely to be different for this group, compared to the non-dormitory clusters and unlinked cases, the only alternative explanation is a higher value of *r* (=2.2).

**Fig. 6.**
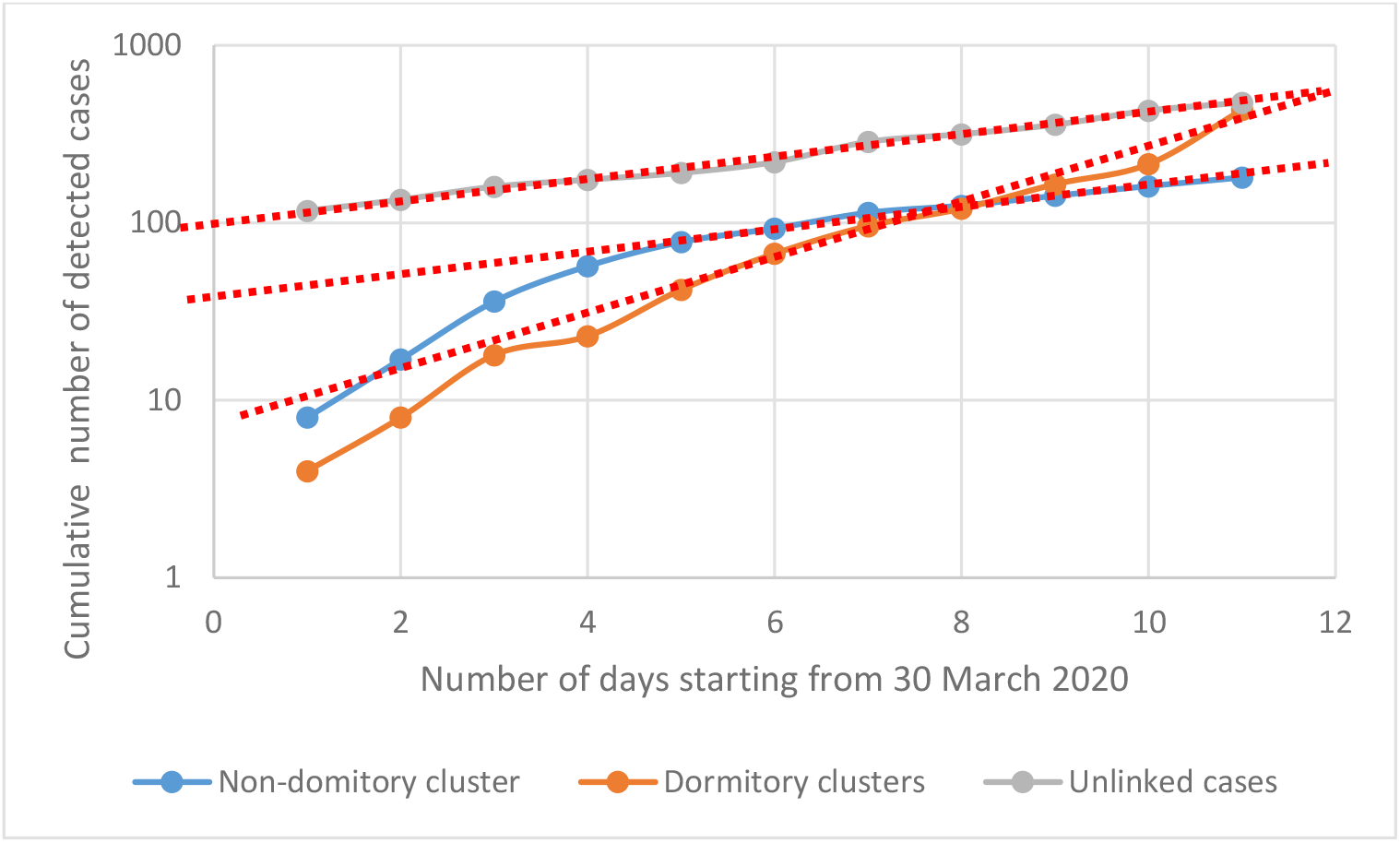
Cumulative cases from dormitory clusters, non-dormitory clusters and unlinked cases starting from 30 March 2020 (data from MOH 2020).

**Fig. 6**. Cumulative cases from dormitory clusters, non-dormitory clusters and unlinked cases starting from 30 March 2020.

## 8 Modelling of Herd Immunity

Herd immunity can be readily incorporated through Eqs. 18 and 19 together with

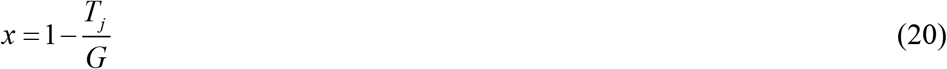

in which *T*_*j*_ is the total number infected up to the point of time, and *G* is the size of the population. Fig. 7 shows the effect of herd immunity on a population size of 5.7 million, with an initial *r* of 1.5, and different values of *k*. As can be seen, the stabilization time and number are both affected by the detection efficiency *k*. With a low detection efficiency, stabilization occurs faster as the epidemic works through the population, but stabilization occurs at a higher total number of infected cases.

**Fig. 7.**
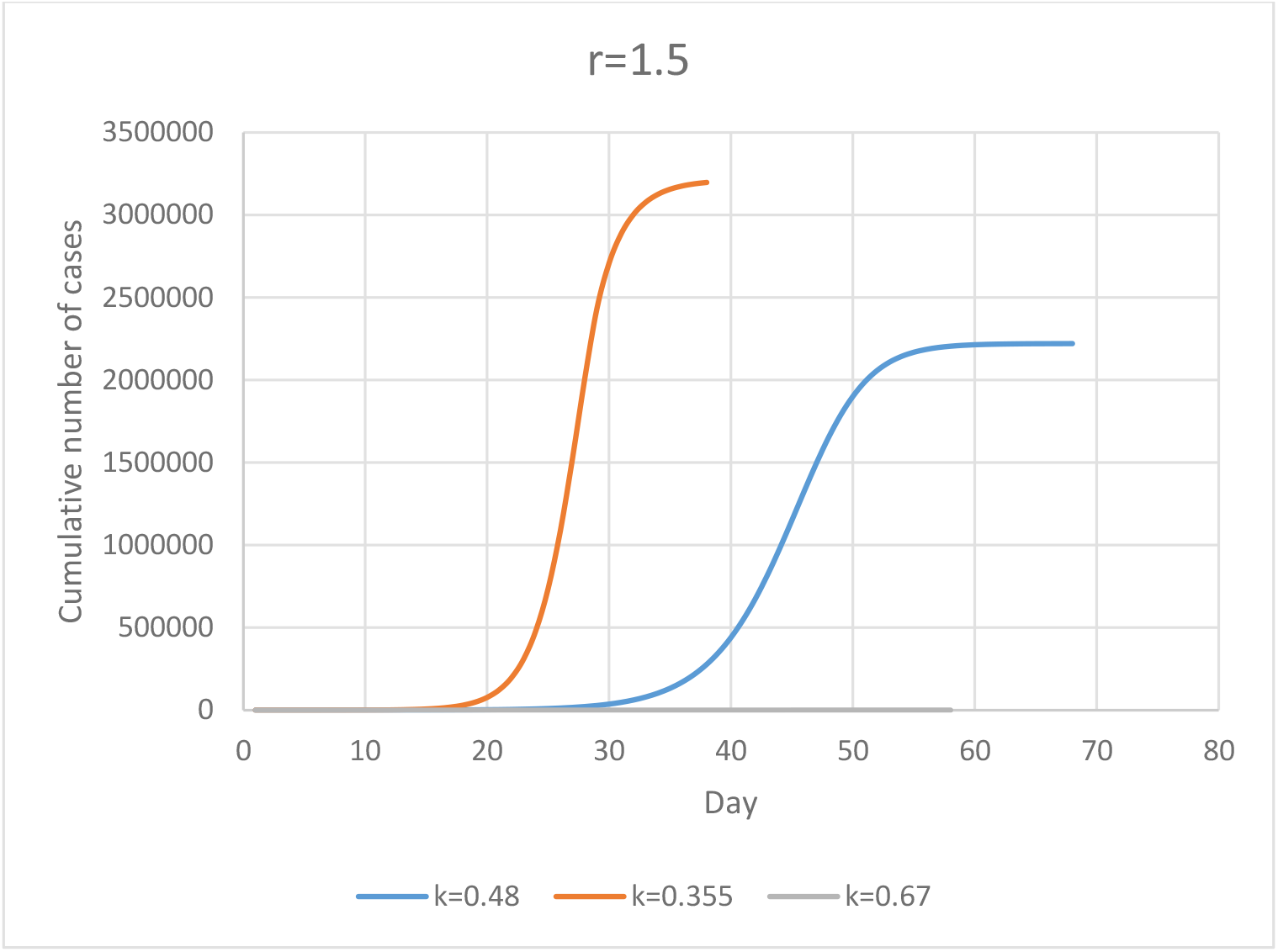
Effect of herd immunity on a population size of 5.7 million.

Modelling of Asymptomatic Spread

The model can be extended to asymptomatic spread, although this requires some re-organization of the spreadsheet, since the detected and undetected cases arising from each day of the asymptomatic period need to be tracked. There is also a possibility that the *k* and *r*-values may differ between asymptomatic and symptomatic periods. To cater for this, separate parameters are used for asymptomatic and symptomatic period; these being designated as *k*_*a*_ and *k*_*s*_, *and r*_*a*_ and *r*_*s*_, where the subscripts *a* and *s* denote asymptomatic and symptomatic parameter. Fig. 8 shows the effect of asymptomatic period on the overall spreading for the values of *k* and *r* as shown in Table 2. The main effect of a low asymptomatic detection efficiency is a slower pick-up in the initial number of detected cases; but this is more than offset by a faster pick-up as the larger number of symptomatic cases kicks in. Not much is known about the asymptomatic detection efficiency but one may surmise that this should be lower than the symptomatic value. The asymptomatic daily infectivity is also highly debated. As more information comes to light in future, these parameters may be more reliably estimated.

**Table 2.** Parameters for Cases 1 to 3 with asymptomatic periods

| Case | Asymptomatic |  | Symptomatic |  |
| --- | --- | --- | --- | --- |
| | Detection efficient $k_a$ | Daily infectivity $r_a$ | Detection efficient $k_s$ | Daily infectivity $r_s$ |
| 1 | 0.3 | 0.5 | 0.5 | 1.5 |
| 2 | 0.3 | 0.6 | 0.5 | 1.5 |
| 3 | 0.1 | 0.6 | 0.5 | 1.5 |

<sup>1</sup> National University of Singapore (non-medical researcher)

**Fig. 8.**
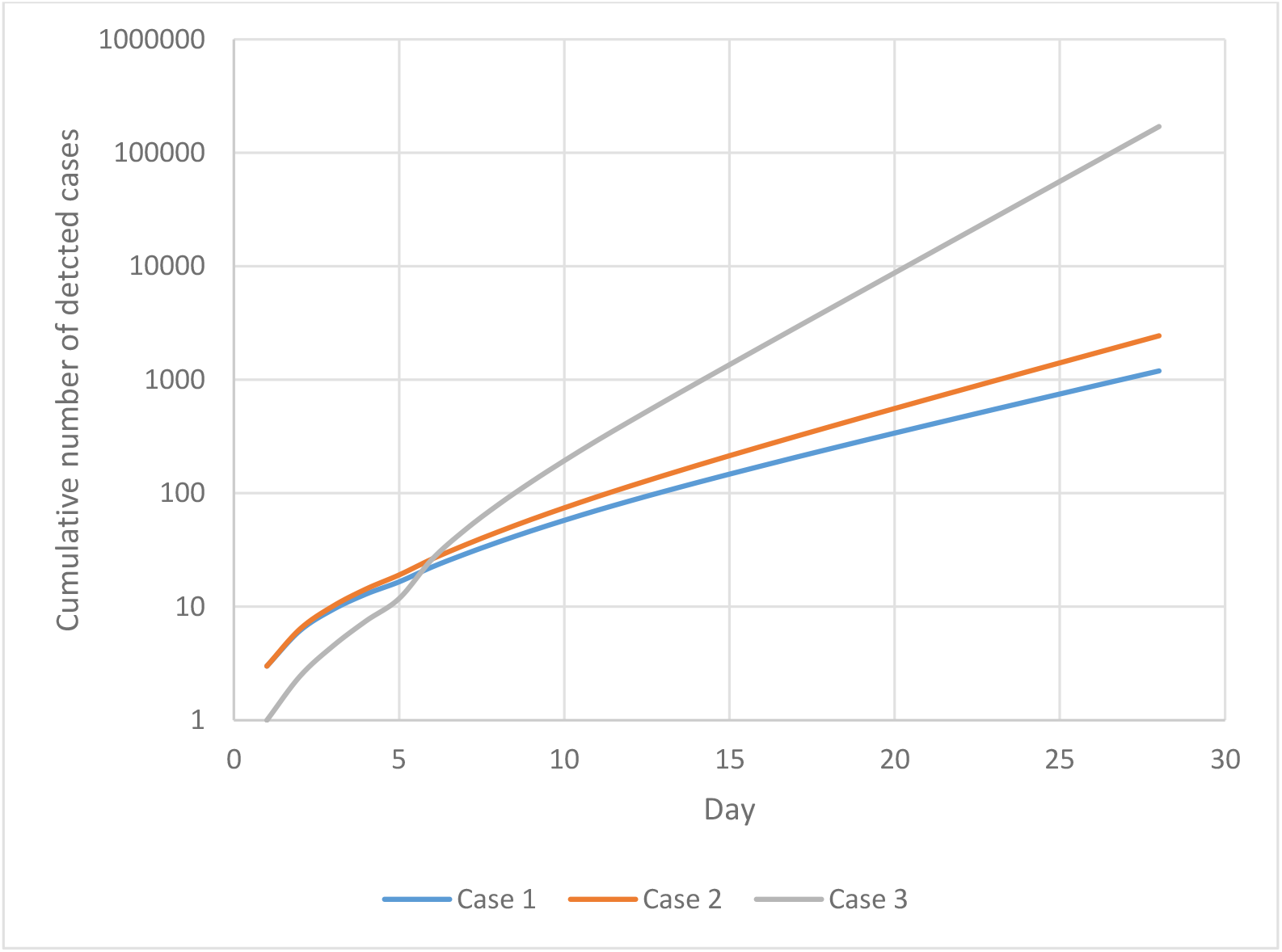
Effect of asymptomatic period on overall spread for different cases, with parameters as shown in Table 2.

## 9 Discussion

An important feature of this model is its simplicity. It can be readily implemented on a spreadsheet or any other programming environment, and therefore potentially widely accessible. There are only two parameters in the models although these parameters may be not time-invariant. The detection efficiency, for instance, may changes as contact tracing and testing are ramped up or down. Similarly, the daily infectivity may also be function of the community setting. The fact that these two parameters can be related to real changes in conditions is an advantage since they allow the effect of any changes in conditions to quantified. As the body of data on the relationship between these parameters and real conditions grows, the model will become increasingly reliable and definitive.

The detection efficiency is not affected by undetected case because back-fitting is based on the rate of increase in the number of detected (or reported) cases rather than the total number of infected cases. In fact, as illustrated in the examples above, if the daily infectivity can be independently estimated, the detection efficiency can be estimated through back-fitting the rate of increase in the number of detected cases.

It can also be readily up-scaled and adapted to different scenarios and conditions. This is especially true for the spreadsheet implementation. As such, this model may be useful as a tool for scenario planning and forecasting not just for an entire population, but also for a specific community within a population. For instance, back-fitting of the data suggests a higher daily infectivity for the dormitory clusters than for the other clusters.

## Data Availability

All data in the manuscripts were obtained from public domain. There are no proprietary data.

